# More Shots on Goal: Heterologous Influenza Prime-Boost Vaccination Decreases the Likelihood of a Negligible Antibody Response to Current or Historical Variants

**DOI:** 10.64898/2026.10.28.26366744

**Authors:** Sierra Orsinelli-Rivers, Ted M. Ross, Tal Einav

## Abstract

Seasonal influenza vaccination only elicits a strong and durable antibody response in a subset of recipients, and prior homologous booster trials have failed to improve on single-dose vaccination. Here, we conducted a phase 4 clinical trial to test if a heterologous prime-boost increases antibody magnitude or breadth as well as reduces the rate of non-responders (ClinicalTrials.gov ID# <u>NCT06567860</u>). Participants received a late-season 2023 inactivated influenza vaccine followed by the 2024 booster four months later containing an updated H3N2 strain alongside unchanged H1N1 and B/Victoria strains, allowing within-subject comparison of heterologous and homologous boosting. The booster vaccination halted or reversed antibody titer waning for all vaccine strains, most notably restoring seroprotective and seroconverting titers for B/Victoria at the cohort-level. Critically, the heterologous booster decreased the rate of non-responders (exhibiting HAI≤10 at all time points), with 0% H3N2 non-responders among subjects receiving the prime and boost compared to 29% H1N1 non-responders. A similar pattern held in a larger cohort administering consecutive fall vaccinations, where non-response rate was 20% of H3N2 compared to 37% for H1N1. Across all non-responders that had consistently low titers against the vaccine strains, up to half elicited an antibody response against historical variants, suggesting that immune imprinting partially underlies the non-responder phenotype. Despite differing vaccination timing, late-season priming and standard fall vaccination produced comparable antibody breadth 30 days post-vaccination when cohorts were matched on pre-vaccination titers. These findings suggest that heterologous vaccinations or other multi-strain exposures can reduce the fraction of the population that does not mount an antibody response during an influenza season.

## Introduction

Seasonal influenza vaccination remains the most effective public health strategy to reduce morbidity and mortality, yet immune responses vary widely across the population. In multi-season analyses spanning 2014-2022, roughly 67% of vaccinees mounted little-to-no antibody response as measured by hemagglutination inhibition (HAI), with their maximum fold-change in antibody titers remaining <4x at all time points.^1–4^ In contrast, only a small fraction of individuals mounted a strong and durable antibody or B cell response, with ∼16% of the population maintaining antibody fold-change≥4x out to 1-year post-vaccination.^3,5^ Longitudinal studies show that most individuals alternate between strong and weak responses across seasons,^6^ suggesting that individuals are primed to respond better to specific influenza antigens. However, the mechanisms underlying this non-responder phenotype remain poorly understood.

To enhance the antibody response and prevent the non-responder phenotype, prior clinical studies have tested the effect of an influenza booster administering an additional dose. Nearly all such studies examined a *homologous booster* using the same influenza vaccine strains administered ∼1 month after the priming dose. These studies often found little-to-no additional increase in HAI titer, and in some cases reduced post-vaccination titers and T cell responses compared to single vaccination.^7–14^ Recently a first-in-class *heterologous booster* was examined that administered the 2018 influenza priming vaccine at the end of the season followed by the 2019 booster vaccine (with different H1N1 and H3N2 strains) three months later, thereby using updates to the seasonal vaccines (which expire on June 30 of each year, marking the end of the season) as the means to administer different strains.^15^ This heterologous booster markedly increased titers against the updated 2019 H3N2 strain (fold-change=3.3-7.1x 1-month post-boost), although the fraction of non-responders was not quantified. Moreover, the heterologous booster minimally increased the antibody response against the 2018 H1N1, 2018 H3N2, or the updated 2019 H1N1 vaccine strains (fold-change≤2x),^15^ suggesting a partial benefit from the heterologous vaccine composition, even at the cohort-level.

This work reexamines a heterologous prime-boost during the 2023-2024 seasons using an egg-grown inactivated influenza vaccine. These two seasons were marked by several notable features. First, the H3N2 vaccine strain was updated from 2023→2024 (H3N2 A/Darwin/9/2021 → A/Massachusetts/18/2022), providing a heterologous booster. Second, the H1N1 and B/Victoria strains did not change (H1N1 A/Victoria/4897/2022, Vic B/Austria/1359417/2021), providing a homologous booster comparison. Lastly, the influenza vaccine shifted from quadrivalent in 2023 to trivalent in 2024 following the removal of the extinct B/Yamagata lineage, which could further improve the antibody response to the remaining three vaccine strains.

Following both the prime and boost, antibody responses were measured against a panel of 17 historical influenza variants for 3-6 time points to assess the magnitude, quality (*i.e.*, breadth), and durability of the antibody response, as well as the fraction of non-responders. For comparison, these late-season vaccine dynamics were assessed against a separate cohort receiving the standard 2023 and 2024 influenza vaccines each fall.

## Results

### Overview of the Cohort

We established a cohort to: 1) compare late-season influenza vaccination in May 2024 to standard vaccination during the fall of 2023 and 2) assess the effects of an influenza booster vaccine 4 months (fall of 2024) after the late-season priming vaccine, henceforth referred to as *prime* and *booster* vaccinations, respectively. This was a phase 4 clinical trial, where 50 participants were screened, 23 subjects were enrolled to receive the late-season prime, of which 14 subjects also received the booster vaccine four months later while the other 9 subjects finished the study and had no more blood draws (**Figure 1**, **Table 1**). All influenza vaccines administered used standard-dose Fluzone, with serum collected 0, 30, and 90 days after each vaccination and measured by HAI against a panel of 17 influenza variants including the 2023-2024 vaccine strains (**Table 2**). All subjects were 18-64 years old, were in good health, and met all eligibility criteria (full details at <u>NCT06567860</u>). The study was conducted at the La Jolla Institute for Immunology (LJI) from May 2024 to February 2025 and approved by the La Jolla Institute for Immunology Review Board (IRB#: VD-286). All participants gave written informed consent prior to enrolling.

**Figure 1.**
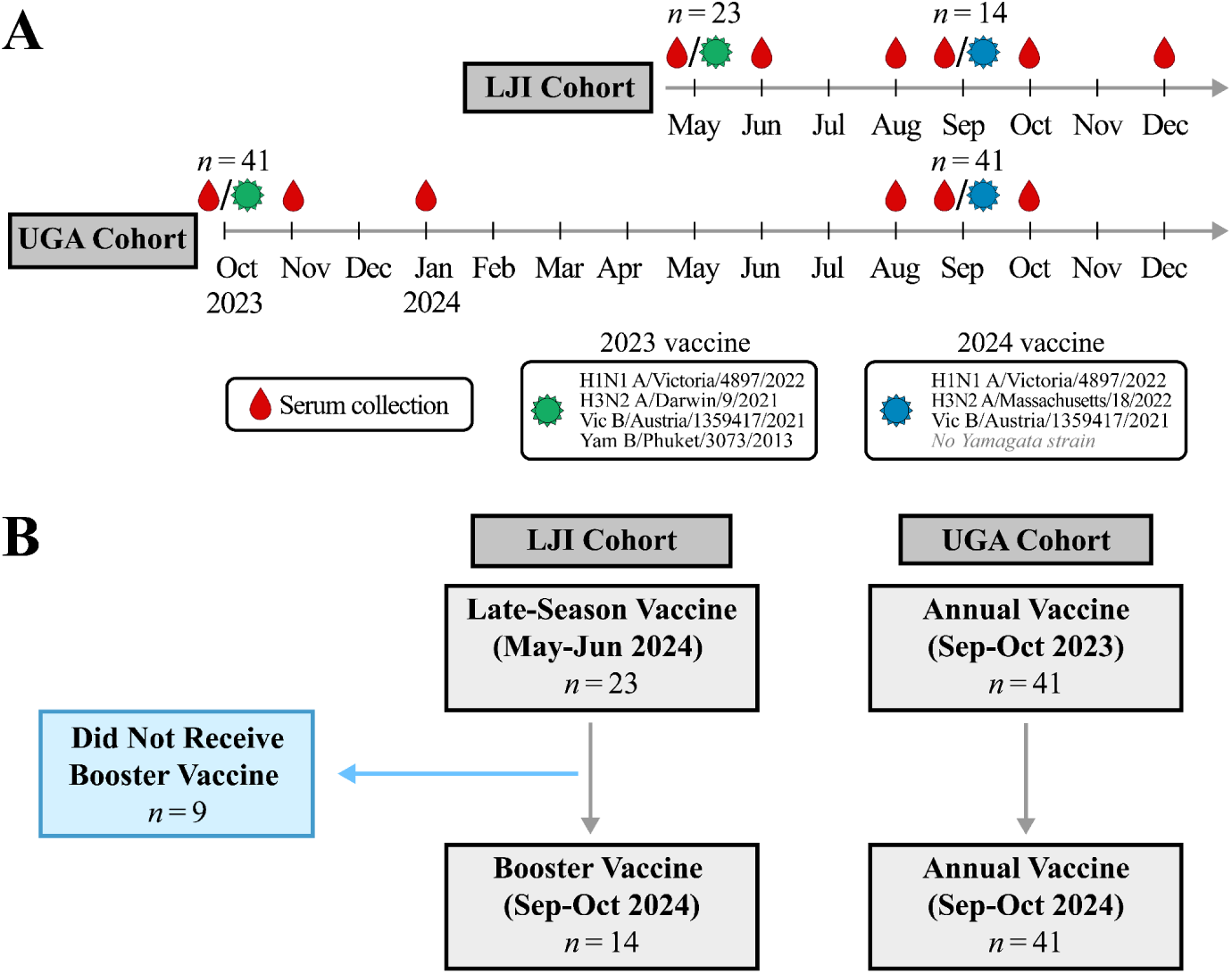
Study design for the LJI and UGA cohorts. (A) Timeline of vaccination and serum collection. (B) Flow diagram tracking subject’s continued participation in both study types.

**Table 1.** Demographics of cohorts analyzed. Demographics for the 23 LJI subjects given the late-season vaccine (2023 LJI) and the subset of 14 subjects that subsequently received the 2024 vaccine during the following fall (2024 LJI). The same 41 UGA subjects were given both the 2023 and 2024 vaccine in the fall. Percentages use the total number of participants in each cohort.

| Variable | Statistic | 2023 LJI<br>(n=23) | 2024 LJI<br>(n=14) | 2023 UGA<br>(n=41) | 2024 UGA<br>(n=41) |
| --- | --- | --- | --- | --- | --- |
| Age (years) |  |  |  |  |  |
|  | Mean (SD) | 29.5 (6.8) | 28.4 (5.2) | 60.3 (17.6) | 61.3 (17.6) |
|  | Min–Max | 22.0–52.0 | 22.0–38.0 | 18.0–88.0 | 19.0–89.0 |
| Sex |  |  |  |  |  |
| Female | n (%) | 13 (56.5) | 8 (57.1) | 25 (61.0) | 25 (61.0) |
| Male | n (%) | 10 (43.5) | 6 (42.9) | 16 (39.0) | 16 (39.0) |
| Vaccine Dose |  |  |  |  |  |
| High dose | n (%) | 0 (0.0) | 0 (0.0) | 22 (53.7) | 24 (58.5) |
| Standard | n (%) | 23 (100.0) | 14 (100.0) | 19 (46.3) | 17 (41.5) |
| Years since Last Influenza Vaccine |  |  |  |  |  |
| <1 years ago | n (%) | 0 (0.0) | 14 (100.0) | 0 (0.0) | 0 (0.0) |
| 1–1.9 years ago | n (%) | 2 (8.7) | 0 (0.0) | 39 (95.1) | 41 (100.0) |
| 2–2.9 years ago | n (%) | 8 (34.8) | 0 (0.0) | 2 (4.9) | 0 (0.0) |
| ≥3 years ago | n (%) | 3 (13.0) | 0 (0.0) | 0 (0.0) | 0 (0.0) |
| Never | n (%) | 8 (34.8) | 0 (0.0) | 0 (0.0) | 0 (0.0) |
| Unknown | n (%) | 2 (8.7) | 0 (0.0) | 0 (0.0) | 0 (0.0) |

**Table 2.** Influenza variants measured by HAI in both cohorts. Vaccine strains are *italicized,* and their background color denotes that they were in the 2023 vaccine only (green; H3N2 A/Darwin/9/2021 and Yam B/Phuket/3073/2013), 2024 vaccine only (blue; H3N2 A/Massachusetts/18/2022), or in both years (purple; H1N1 A/Victoria/4897/2022 and Vic B/Austria/1359417/2021). The Yamagata B/Phuket/3073/2013 strain was included in the 2023 vaccine but removed from the 2024 vaccine due to lineage extinction.

| Influenza A, H3N2 | Influenza A, H1N1 | Influenza B, Victoria | Influenza B, Yamagata |
| --- | --- | --- | --- |
| A/Hong Kong/4801/2014 | A/California/07/2009 | B/Colorado/6/2017 | *B/Phuket/3073/2013* |
| A/Singapore/INFIMH-16-00<br>19/2016 | A/Brisbane/2/2018 | B/Washington/2/2019 |  |
| A/Kansas/14/2017 | A/Guangdong-Maonan/S<br>WL1536/2019 | *B/Austria/1359417/2021* |  |
| A/South Australia/34/2019 | A/Victoria/2570/2019 |  |  |
| A/Hong Kong/2671/2019 | *A/Victoria/4897/2022* |  |  |
| A/Tasmania/503/2020 |  |  |  |
| *A/Darwin/9/2021* |  |  |  |
| *A/Massachusetts/18/2022* |  |  |  |

Antibody titers in the LJI cohort were compared against standard influenza vaccination administered each fall within the annual University of Georgia (UGA) cohort, where *n*=41 individuals with age 18-90 were given both the 2023 influenza vaccine in the fall of 2023 and the 2024 vaccine in the fall of 2024. The study design was standardized as much as possible to the LJI study for both the time points measured and influenza variants assessed by HAI, although day 90 was not collected post-2024-vaccination and 3 of the 17 variants could not be measured in a subset of the post-2023-vaccination time points (**Methods**). Despite these similar protocols, UGA subjects tended to be older and more recently vaccinated, which could affect antibody responses (**Table 1**).^16^ All subjects were given either Fluzone (age<65) or Fluzone High Dose (age≥65). Details on the longitudinal UGA cohort have been described previously.^4,17^

### Influenza Prime-Boost Vaccination led to Durably Elevated Antibody Titers, with Fewer Non-Responders for Heterologous H3N2 than Homologous H1N1

In the 2023 LJI cohort, the late-season priming vaccination elicited peak HAI fold-change (titer relative to pre-vaccination) of 3.9-6.7x across all vaccine strains 30 days post-vaccination (**Fig 2**). Fold-change then decreased to 2.3-4.7x at day 90 and 1.7-3.5x at day 120, following the kinetics of standard fall vaccination seen in multiple prior studies.^3^ The vaccine strains for H1N1, H3N2, and B/Victoria maintained significantly higher geometric mean titers (GMTs) at every post-vaccination time point compared to pre-vaccination GMTs at day 0 (adjusted *p* ranging from <0.001 to 0.015, one-sided Welch’s t-test with Holm’s correction used throughout analysis, **Table S1**). Interestingly, the H1N1, H3N2, and B/Victoria vaccine strains all had pre-vaccination GMT<10, whereas B/Yamagata had substantially elevated pre-vaccination GMT=47 and maintained these higher titers at all time points.

**Figure 2.**
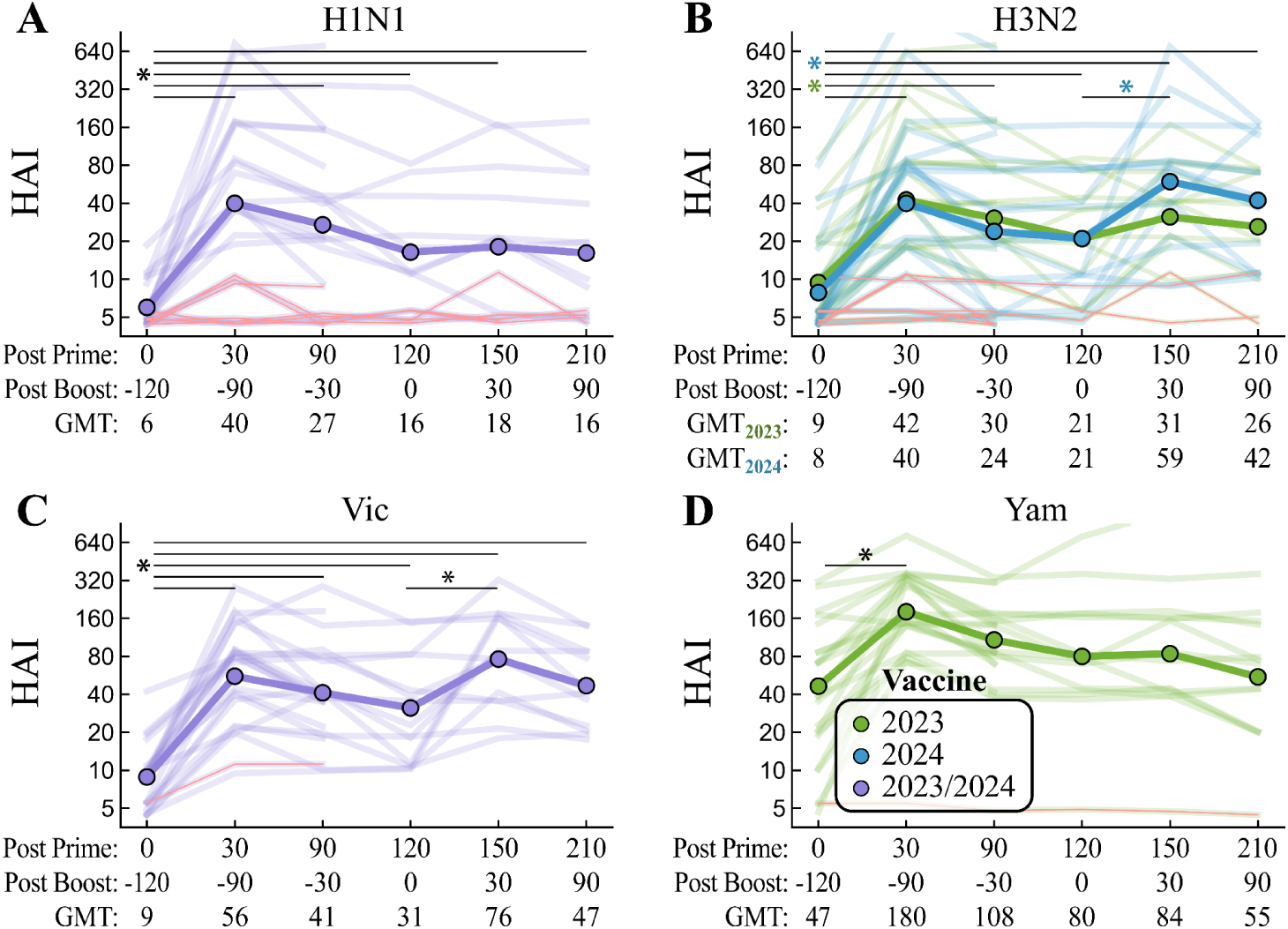
Longitudinal HAI titers for vaccine strains after influenza prime and boost vaccinations. Titers for the (A) H1N1, (B) H3N2, (C) B/Victoria, or (D) B/Yamagata vaccine strains from the 2023-2024 LJI cohort. Plot markers represent GMTs, with individual subjects shown with opacity. Strains only in the 2023 vaccine are colored in green (H3N2 A/Darwin/9/2021, Yam B/Phuket/3073/2013), those only in the 2024 vaccine are colored in blue (H3N2 A/Massachusetts/18/2022), and strains present in both years are colored in purple (H1N1 A/Victoria/4897/2022 and Vic B/Austria/1359417/2021). There was no Yamagata strain in the 2024 vaccine. Red lines denote subjects with HAI titer≤10 at all time points. *: *p*≤0.05 (Welch’s one-sided t-test on log-transformed titers with Holm correction) across time points; all *p*-values listed in **Table S1**. The two colored asterisks in Panel B denote that both the 2023 and 2024 vaccine strains showed significantly higher GMTs at every post-vaccination time point, while titers only significantly increased against the 2024 vaccine strain from day 120 to day 150.

The 2024 booster vaccination either halted or reversed the decrease in titers from days 30→120, with H1N1 maintaining its GMT titers at 150 days post-prime (30 days post-boost) while GMTs to 2024 H3N2 and B/Victoria significantly increased by 1.5x (adjusted *p*=0.031) and 2.8x (adjusted *p*=0.017) compared to 120 days post-prime, respectively (**Fig 2**). Titers for all three vaccine strains showed slight-to-moderate decay by 210 days post-prime (90 days post-boost), with H1N1 having the lowest GMT=16 (fold-change=2.7x relative to pre-2023-vac) while H3N2 and B/Victoria had GMT=42-47 (fold-change=5.3-5.4x relative to pre-2023-vac).

The 2024 booster vaccine did not contain B/Yamagata, and this lineage’s GMT trajectory followed the expected decay of a single influenza vaccination with titers decaying to ≈1.5x of the pre-vaccination level.

Surprisingly, 30%=7/23 subjects were non-responders for H1N1, maintaining HAI titers≤10 at all time points, compared to only 4%=1/23 subject for B/Victoria and B/Yamagata (**Fig 2**, red lines). The H3N2 vaccine strain changed from A/Darwin/9/2021 in 2023 to A/Massachusetts/18/2022 in 2024, and none of the subjects (0/14) that received the prime and boost had HAI titers≤10 against both vaccine strains at all time points. 22%=2/9 subjects [ID=4042, 4937] that only received the prime boost had titers≤10 to both strains at days 0, 30, and 90, but neither subject received the 2024 vaccine and hence did not have the opportunity to respond to H3N2 A/Massachusetts/18/2022 (individual subjects shown in **Figs S1**). An additional 2 subjects [ID=4884, 4932] maintained titers≤10 for the 2023 vaccine alone and another 2 subjects [ID=4880, 4936] achieved this for the 2024 vaccine alone. In short, by providing two H3N2 antigens, the heterologous prime-boost decreased the likelihood that a subject maintained uniformly low antibody titers against both vaccine strains.

### Prime-Boost Elicited Substantial Breadth to H1N1 Variants, with several Non-Responders to the Vaccine showing an Antibody Response to Historical Variants

HAI titers were measured against a panel of 17 influenza variants spanning multiple decades to evaluate whether late-season priming and subsequent boosting broadened the antibody repertoire both at the cohort- and subject-level (**Table 2**). At the 2023 baseline (day 0), the H1N1, H3N2, and B/Victoria vaccine strains exhibited the lowest GMTs among their respective variant groups, even though the same H3N2 and B/Victoria vaccine strains were also included in the prior 2022 influenza vaccine (**Fig 3A**).

**Figure 3.**
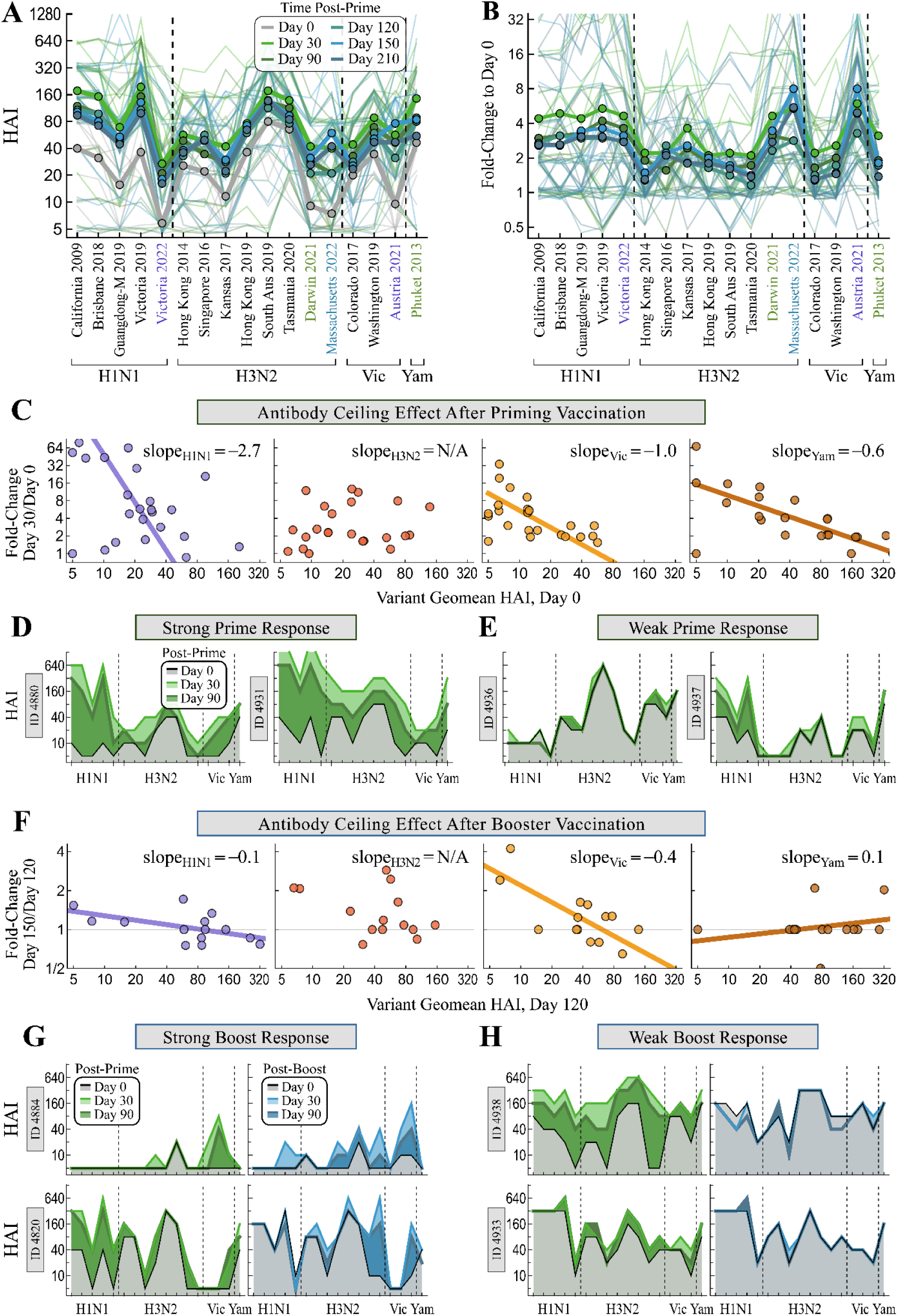
Antibody breadth post-prime and post-boost vaccination against a panel of influenza variants. (A) HAI titer and (B) fold-change relative to day 0 (priming vaccine) for LJI subjects that received both the 2023 and 2024 vaccines. (C) Linear fits of fold-change for each subject 30 days after priming vaccination vs pre-prime GMTs against all variants, both on logged axes. Antibody landscapes for two individuals with (D) strong or (E) weak antibody responses following the priming vaccine (all subjects in **Fig S1**). (F) Linear fits of fold-change for each subject 30 days after booster vaccination vs pre-boost (day 120 post-prime) GMTs against all variants. Antibody landscapes for two individuals with (G) strong or (H) weak antibody booster responses following the priming vaccine. In Panels C/F, H3N2 showed weak correlation and was not fit. In Panels D/E/G/H, variants on the x-axis are shown in the same order as in Panels A/B.

The breadth of the response following prime and boost differed across each subtype/lineage, with all H1N1 variants exhibiting GMT fold-change>4x at 30 days post-prime, whereas only the 2023 and 2024 H3N2 and B/Victoria vaccine strains did so (**Fig 3B**). Although titers declined at later time points, H1N1 maintained a larger mean fold-change≈3x across all prior variants at days 120-210 post-prime compared to ≈1.7x for H3N2 and 1.4x for B/Victoria. The booster vaccination at day 120 post-prime elicited a small, transient increase in titers for prior H1N1 and H3N2 variants but no increase for prior B/Victoria variants.

The antibody ceiling effect, where individuals with larger pre-vaccination titers tend to exhibit smaller fold-change post-vaccination,^18,19^ was tightly observed in both influenza B lineages, where 2.0x (Vic) or 3.1x (Yam) larger pre-vac GMTs against all variants resulted in 2x smaller fold-change across all subjects (**Fig 3C, Methods**). As a result, individuals with larger pre-vac titers usually exhibited smaller fold-change but still ended up with larger post-vaccination titers.

H1N1 exhibited a stronger antibody ceiling where 1.3x larger pre-vac GMTs led to 2x smaller fold-change, although multiple subjects broke this trend. For H3N2, pre-vac GMTs and fold-change were weakly correlated and no ceiling effect was observed. Overall, while individuals with small pre-vac titers often had large fold-change (**Fig 3D**), a few subjects with small pre-vac titers responded weakly to the priming vaccine (**Fig 3E**, left; all LJI subjects shown in **Fig S1**).

Following the booster vaccination, both the fold-change and antibody ceiling effect (relative to day 120) were smaller for each vaccine strain. B/Victoria showed the steepest slope, where a 5.0x larger pre-booster GMT resulted in 2x smaller fold-change (**Fig 3F**). H1N1 and B/Yamagata had nearly flat slopes suggesting no ceiling, while H3N2 still showed weak correlation with no clear ceiling effect.

At the individual-level, some non-responders (categorized using their HAI to the vaccine strains) exhibited an antibody response against prior variants. For H1N1, 2/7=29% of subjects [ID=4884, 4932] showed fold-change≥4x (seroconversion) to an older H1N1 variant (**Fig S1**). In fact, ID=4884 was the only subject in the LJI cohort with HAI=5 against all H1N1 variants at day 0, yet they showed no H1N1 response to the priming vaccine and a transient response at day 30 post-boost that was entirely gone by day 90 post-boost (**Fig 3G**, top). While their B/Yamagata titers remained at HAI=5 across all time points, their B/Victoria titers increased nearly identically post-prime and post-boost. Finally, their H3N2 prime response was weak and transient, while their booster response was far stronger, broader, and more durable, demonstrating that the updated 2024 H3N2 vaccine strain elicited a better response in this individual.

Individual longitudinal trajectories showed that subjects with the strongest responses to 2nd vaccination tended to have transient prime responses with antibody titers at day 120 (day booster administered) similar to their day 0 titers (day prime administered) (**Fig 3G**, bottom). In contrast, subjects whose antibody titers remained elevated at day 120 or whose pre-vac titers were very high responded weakly to 2nd vaccination (**Fig 3H**). While a subject’s antibody responses to the homologous 2024 H1N1 and B/Victoria vaccine strains was either comparable or weaker than their response to the 2023 vaccine, a few subjects showed stronger responses to the 2024 H3N2 responses compared to the 2023 vaccine. Collectively, these results suggest that a homologous booster can reelevate transient antibody responses, and that a heterologous H3N2 booster can elicit enhanced responses.

## Standard Fall Vaccination showed Similar Kinetics but weaker H1N1 Responses

To determine whether late-season priming vaccination alters immune kinetics compared to standard fall vaccination, we next evaluated the dynamics of the n=41 individuals in the 2023-2024 UGA cohort (**Fig 4**). Serum was collected at days 0, 30, and 90 post-2023-vaccination, as well as day 365 (pre-2024 dose) and day 395 (30 days post-2024-vaccination). However, HAIs to the H1N1 vaccine strain were not measured at day 90, nor were H3N2 A/Hong Kong/4801/2014 and H3N2 A/Massachusetts/18/2022 (the 2024 vaccine strain) measured at day 0, 30, or 90.

**Figure 4.**
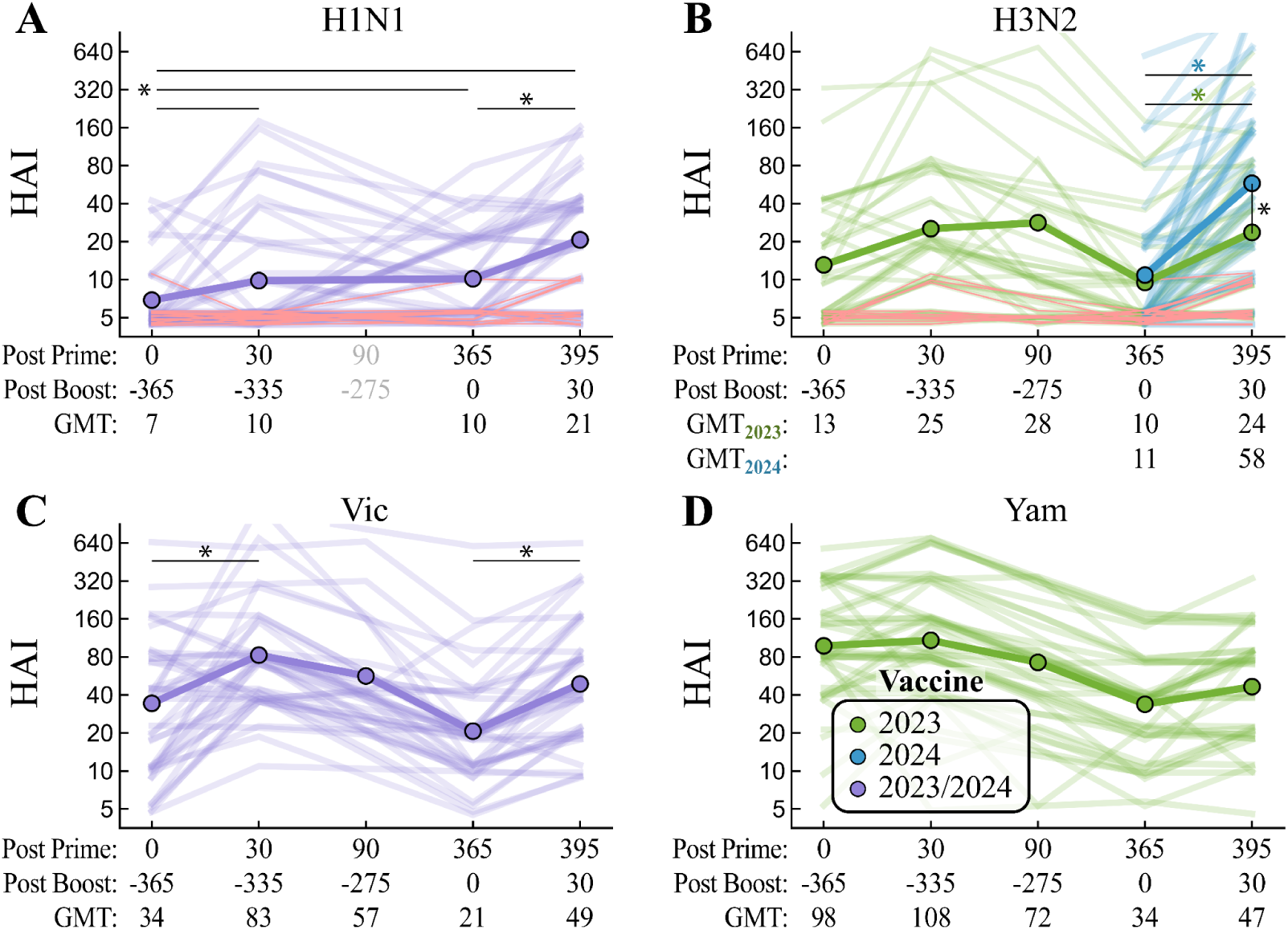
Longitudinal HAI titers for vaccine strains after two consecutive fall vaccinations. Titers for the (A) H1N1, (B) H3N2, (C) B/Victoria, or (D) B/Yamagata vaccine strains from the 2023-2024 UGA cohort, analogous to Fig 2. Red lines denote subjects with HAI titer≤10 at all time points. *: *p*≤0.05 (one-sided t-test on log-transformed titers with Holm correction), with horizontal bars comparing time points and vertical bars (for H3N2) comparing the 2024 and 2023 vaccine strains; all *p*-values listed in **Table S2**. The green and blue asterisks in Panel B represent significance for the 2023 and 2024 H3N2 vaccine strains, respectively.

Following 2023 vaccination, H1N1 post-vaccination titers were surprisingly low in the UGA study, with the subsequent GMT=10 on day 30 representing a small but significant increase (adjusted *p*=0.040) despite this fold-change being markedly smaller than what was observed in the LJI cohort (fold-change_UGA,H1N1_=1.4x, fold-change_LJI,H1N1_=6.7x) (**Fig 4**). GMTs for H3N2 increased (adjusted *p*=0.071) and B/Victoria significantly increased at day 30 (adjusted *p*=0.006) and stayed within 2-fold at day 90. B/Yamagata began with a high baseline GMT=98 that did not significantly increase at day 30 (adjusted *p*=1.0) before slightly decreasing at day 90.

Prior to the 2024 vaccination at day 365, GMTs were 0.4‒0.7-fold as large in the UGA cohort compared to the LJI cohort at day 120, likely because of the longer UGA waning period.

Between day 365 and day 395, GMTs in the UGA cohort significantly increased for all three vaccine strains (adjusted *p*<0.001 for all) but not the B/Yamagata strain that was no longer in the vaccine (adjusted *p*=0.074). As a result, GMTs across all vaccine strains were ≤2-fold different between the UGA and LJI cohorts at 30 days post-2024-vaccination.

Notably, there was a substantial difference in the fraction of non-responsive individuals between the cohorts, represented by flat trajectories with HAI titers ≤10 across all time points. Across all UGA subjects, 37%=15/41 maintained HAI titers≤10 at all time points for H1N1 (**Fig 4A**). In contrast, zero subjects were non-responsive to B/Victoria and B/Yamagata (**Fig 4C-D**). For H3N2, 29%=12/41 subjects failed to respond to the 2023 vaccine strain (A/Darwin/9/2021) and 27%=11/41 failed to respond to the 2024 strain (A/Massachusetts/18/2022; **Fig 4B**), representing a higher non-responder fraction than observed in the LJI cohort. Eight of these subjects overlapped, failing to respond to either the 2023 or 2024 H3N2 vaccine strains.

Furthermore, six subjects failed to respond to the H1N1 and H3N2 strains both years, demonstrating a persistently poor response to influenza A antigens across both seasons.

### Approximately Half of Non-Responders to Vaccine Strains Elicited an Antibody Response to Historical Variants during Consecutive Fall Vaccinations

We next assessed if antibody breadth was more restricted in the UGA cohort using the panel of 17 influenza variants (**Fig 5A-B**, **Table 2**). Following 2023 vaccination, GMTs minimally increased by roughly 1-2x across all variants at day 30 before decaying to or even below baseline (fold-change≤2x for all strains by day 365). After the 2024 vaccine administered at day 365, all H1N1 and B/Victoria strains at day 395 exhibited fold-change≈2x. H3N2 fold-change was greatest for the updated H3N2 A/Massachusetts/18/2022 vaccine strain (fold-change=5.3x), with smaller fold-change seen for older H3N2 variants.

**Figure 5.**
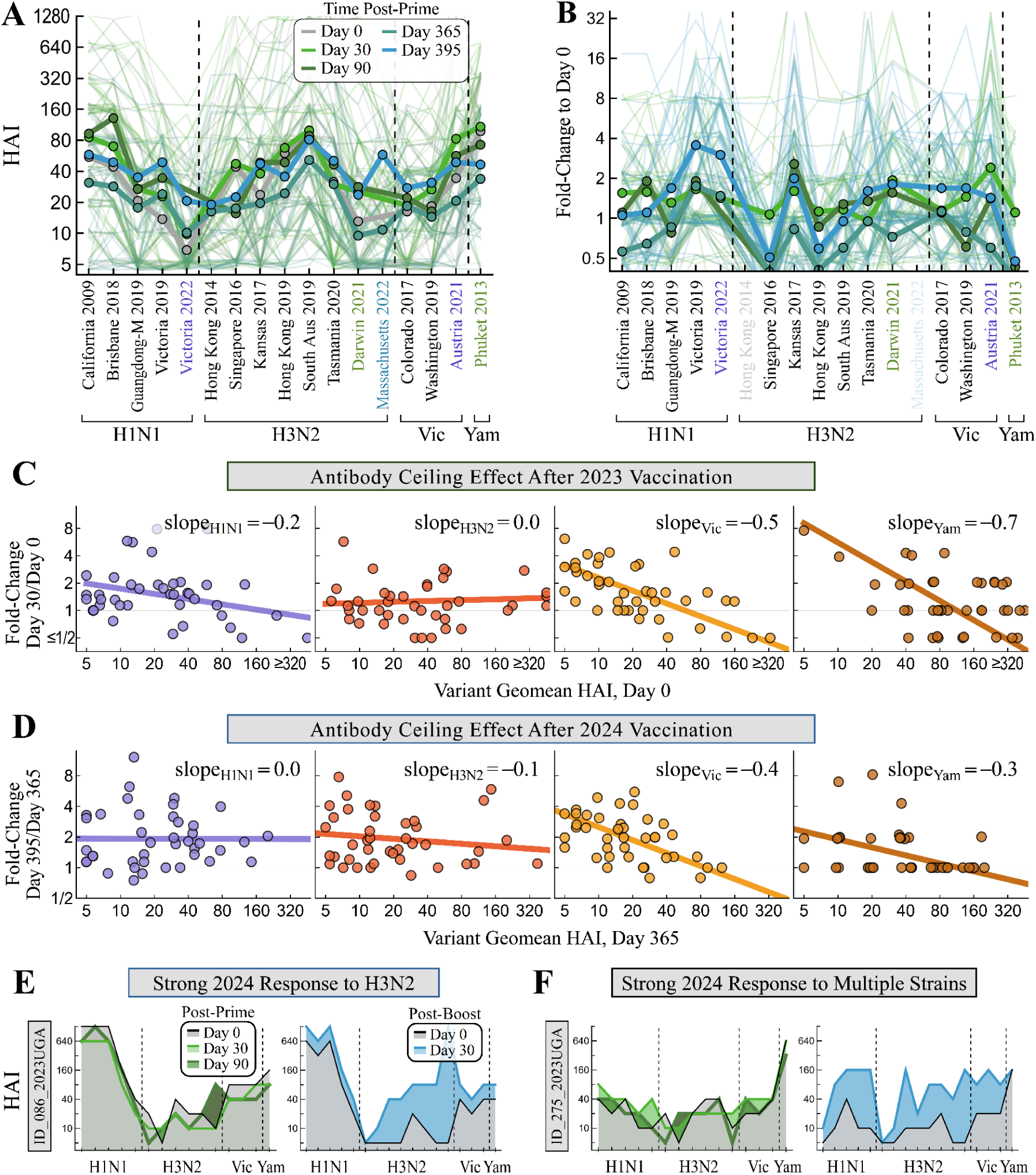
Antibody breadth following two consecutive fall vaccinations against a panel of influenza variants. (A) HAI titer and (B) fold-change relative to day 0 (priming vaccine) for 2023-2024 UGA subjects that received both the 2023 and 2024 vaccine. H3N2 Hong Kong 2014 and Massachusetts 2022 were not measured at days 0, 30, 90 and hence are not shown in Panel B; H1N1 Victoria 2022 was not measured at day 90. Linear fits of fold-change for each subject 30 days after (C) 2023 or (D) 2024 vaccination vs that season’s pre-vac GMTs against all variants, both on logged axes. (E,F) Antibody landscapes for two individuals exhibiting weak 2023 vaccine responses but strong responses (E) mainly to H3N2 strains or (F) to multiple vaccine strains (all subjects in **Fig S2**). In Panels E/F, variants on the x-axis are shown in the same order as in Panels A/B.

The UGA cohort exhibited little-to-no antibody ceiling effect for H1N1 or H3N2 (*i.e.* fold-change is nearly independent of pre-vac titer), while showing a stronger ceiling effect for both influenza B viruses. Following 2023 vaccination, an increase of 4.4x (Vic) or 2.7x (Yam) in pre-vac titers led to 2x smaller fold-change (**Fig 5C**). In 2024, an increase of 5.1x (Vic) or 14.3x (Yam) in pre-vac titers led to 2x smaller fold-change, with B/Yamagata likely showing a much flatter ceiling effect due to its removal from the 2024 vaccine (**Fig 5D**).

Individual longitudinal landscapes further highlighted variable antigen responsiveness across consecutive fall vaccinations. A subset of subjects who responded weakly to the initial 2023 vaccination mounted strong responses following the 2024 vaccination that ranged from strain-specific (H3N2 only in **Fig 5E**) to broad responses (**Fig 5F**). Yet the larger UGA cohort demonstrated that roughly half of non-responders (*i.e.*, HAI titers≤10 against the 2023 and 2024 vaccine at all time points) mounted an antibody response to historical variants. More precisely, 7/15=47% of H1N1 non-responders [ID=077, 179, 353, 438, 443, 614, 629] and 4/8=50% of H3N2 non-responders [ID=296, 569, 589, 600] had fold-change≥4x against a prior influenza variant (**Fig S2**).

### Vaccination Timing shapes Post-Vaccination Antibody Breadth more than Vaccine Composition

To test if vaccine composition or the timing of vaccination have a greater impact on antibody breadth, we compared the 2023 LJI and 2024 UGA studies (temporally close, administering different vaccines) as well as the 2023 LJI and 2023 UGA studies (temporally further, administering the same vaccine) at 30 days post-vaccination. In each case, antibody titers across all H1N1, H3N2, or B/Victoria variants were used to assess how many of the subjects that had quantitatively similar antibody breadth pre-vaccination maintained that similarity post-vaccination.

More precisely, out of all pairs of subjects where the first individual came from 2023 LJI while the second came from either 2023 or 2024 UGA, we computed the root-mean-square error (RMSE) between their HAI titers across all variants within one subtype/lineage at day 0 and day 30. The percent of pairs with RMSE≤4x at day 0 that maintained RMSE≤4x at day 30 was used as a proxy for study similarity (**Fig 6, Methods**). For example, 46.0% of pairs with H3N2 RMSE≤4x pre-vaccination maintained RMSE≤4x post-vaccination between the 2023 LJI and 2023 UGA cohorts (**Fig 6A,C**), while 60.3% did so between 2023 LJI and 2024 UGA (**Fig 6C**).

**Figure 6.**
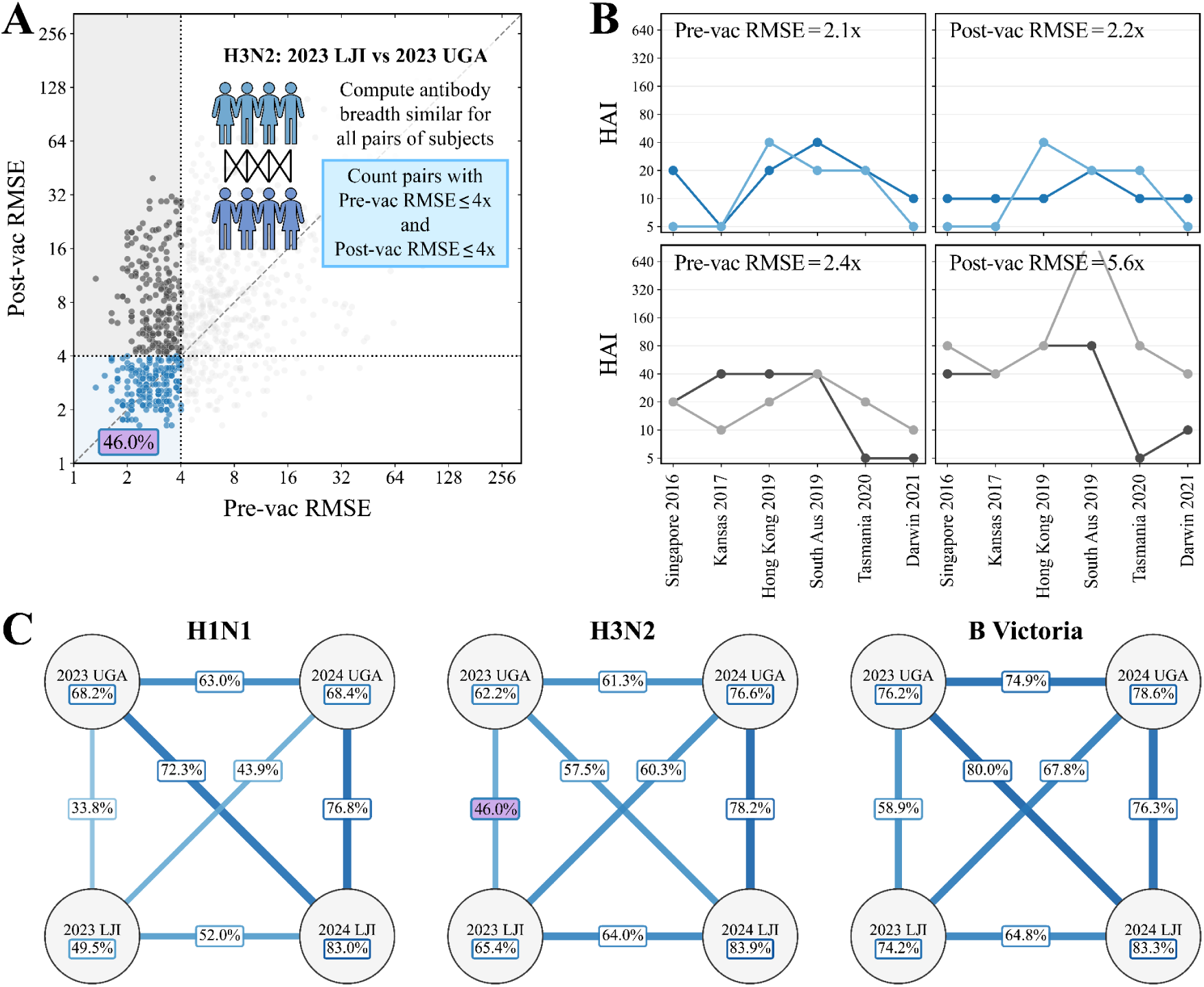
RMSE-based matching of antibody landscapes between LJI and UGA study cohorts. (A) For each pair of subjects between the 2023 LJI and 2023 UGA cohorts, the RMSE across all H3N2 variants is computed pre-vaccination and 30-days post-vaccination. Among subjects with RMSE≤4x pre-vaccination, the fraction that maintains RMSE≤4x post-vaccination serves as a proxy for cohort similarity. (B) Example pair of subjects that maintained RMSE≤4x pre- and post-vaccination and are counted in the numerator and denominator of study similarity (*top plots*, ID=086 from 2023UGA in dark blue and ID=4042 from 2023LJI in light blue). Example pair with RMSE≤4x pre-vac but RMSE>4x post-vac only counted in the denominator of study similarity (*bottom plots*, ID=461 from 2023UGA in dark gray and ID=4910 from 2023LJI in light gray). (C) Generalizing this cohort similarity for H1N1, H3N2, and B/Victoria as well as the 2023-2024 LJI and UGA studies. Lines denote cross-study similarity while circles show within-study similarity. The H3N2 comparison from Panel A is highlighted with a purple background. B/Yamagata was excluded because only a single variant was measured from that lineage.

For H1N1 and B/Victoria, similarity between 2023 LJI and 2024 UGA was also ∼10% higher than between 2023 LJI and 2023 UGA, even though these two vaccine strains did not change, suggesting closer temporal administration of the vaccine can affect the antibody response.

Applying this similarity score between every pair of studies showed that the 2024 LJI and 2024 UGA studies consistently had the largest or second-largest cross-study similarity within each subtype/lineage, which could be partly attributed to all 2024 subjects having been vaccinated during the prior season. Notably, 2024 LJI and 2024 UGA cross-study similarity was generally comparable to the self-similarity of each study (*i.e.*, among pairs of subjects within one study, circles in **Fig 6C**). Overall, these results suggested that while vaccine timing and vaccination history shape antibody breadth, receipt of the 2024 vaccine elicited similar post-vaccination titers whether administered as the culmination of a prime-boost or as two consecutive fall vaccinations.

## Discussion

By administering the 2023 and 2024 influenza vaccines as a prime-boost regiment, this study simultaneously examined the impact of a homologous booster where the vaccine strain did not change (H1N1 and B/Victoria) as well as a heterologous booster (H3N2). At the cohort-level, the booster vaccination in the LJI cohort arrested the decay of antibody titers toward baseline levels as seen in prior studies.^1–4^ Thus, compared to day 0, GMTs remained significantly elevated at every time point through day 210 post-prime (90 days post-boost) for all four vaccine strains.

B/Victoria demonstrated a clear benefit from the LJI booster vaccination, since GMT≥40 and fold-change≥4x relative to day 0 (often called seroprotection and seroconversion) were maintained against the vaccine strain from day 30-90, lost by day 120, and then restored from day 150-210 following the booster. In contrast, H1N1 vaccination did not significantly increase GMTs post-boost. These two vaccine components also had starkly different breadth profiles, with H1N1 titers elevated against all five variants measured (isolated between 2009-2022) while B/Victoria titer increases were focused on the vaccine strain alone. Given that H1N1 shows faster antigenic evolution than B/Victoria,^20,21^ we speculate that this difference in breadth represents a more polyclonal H1N1 antibody response that may underlie why the 2024 booster did not elevate H1N1 titers.

H3N2 GMTs for the 2024 vaccine strain showed similar kinetics to B/Victoria, increasing post-prime and post-boost. Importantly, the two H3N2 antigens gave each subject two opportunities to elicit an H3N2 antibody response. As a result, 0/14 of LJI subjects and 8/41 UGA subjects receiving both the 2023 and 2024 vaccines maintained low antibody titers (HAI≤10 at all time points) against both H3N2 vaccine strains, whereas H1N1 had roughly twice as many non-responders. This suggests that the benefit of heterologous vaccination is that it gives each subject two opportunities to elicit an antibody response. In a season where multiple antigenically distinct H3N2 viruses cocirculate, adding both viruses heterologous prime-boost or in a single vaccine^22^ could increase the likelihood of a protective antibody response against at least one strain.

Among subjects that did not respond to the vaccine strain, 29-50% elicited an antibody response against prior variants, suggesting that this phenotype is partially attributed to imprinting (also known as original antigenic sin), which is potentially shaped by a subject’s infection history.^6,23,24^ However, even children with no prior influenza vaccination or infection history and no measurable HAI titers may not mount an antibody response following inactivated influenza vaccination,^25^ while children receiving their first live-attenuated influenza vaccine are split between mounting a systemic antibody response, mucosal antibody response, or CD8 T cell response.^26^ Future work should investigate whether adult non-responders instead mount a T cell or innate immune response.

This study had several limitations. First, the cohort sizes were small, particularly among the 14 LJI subjects that received both the prime and booster doses, limiting the ability to detect modest effects or stratify by additional variables. Antibody responses were assessed exclusively by HAI to current and prior vaccine strains. Both the LJI and UGA cohorts administered Fluzone, while other vaccine brands can elicit different antibody or cellular responses.^27–29^ In addition, differences between the two cohorts should be interpreted cautiously given that the LJI cohort showed substantially larger H1N1 fold-change than the UGA cohort at day 30 post-2023-vaccination (6.7x vs. 1.4x fold-change). Moreover, UGA participants had higher day 0 GMTs across all four vaccine strains, likely due to differences in prior exposures and the fact that the LJI 2023 priming vaccine happened later in the season than the corresponding UGA priming vaccine, giving more time for antibody waning in LJI subjects. These cohort-level differences suggest that the two populations are not directly comparable, and that some of the discrepancies noted between late-season and standard annual vaccination may reflect underlying cohort differences rather than vaccine timing.

Other types of heterologous boosters are under investigation. Some groups administer both the northern and southern hemisphere vaccines 6 months apart, which may be especially beneficial in countries like Hong Kong that experience two influenza seasons in the winter and summer.^30^ mRNA vaccines can simultaneously administer multiple H3N2 strains to enhance breadth, although initial results suggest that antibody titers were highly localized to each vaccine strain (*i.e.*, the response to one H3N2 strain did not markedly increase or decrease if other H3N2 vaccine strains were included).^22^ Given that we still do not understand which individuals will respond to which strains, and that the response to each vaccine strain appears to be mostly independent of the other vaccine strains, both of these approaches should increase the likelihood of a response to the H1N1, H3N2, and influenza B vaccine components.

Current influenza vaccines present the most common circulating variants, yet not all immune systems react equally well to each virus. Ultimately, frameworks that mechanistically predict each subject’s immunological response to different antigens could prove invaluable in designing vaccines that elicit stronger and more uniform responses across the population.

## Methods

### Study design

The LJI late-season prime-boost study was a Phase 4 clinical trial (<u>NCT06567860</u>) conducted at the La Jolla Institute for Immunology (LJI) from May 2024 to February 2025 (IRB# VD-286). 23 individuals aged 18-64 initially enrolled in this study. Inclusion criteria required adults to be age 18–64 in good general health; exclusion criteria included receipt of a 2023–2024 seasonal influenza vaccine prior to enrollment, history of Guillain-Barré syndrome, egg allergy, active immunocompromising conditions (HIV, HBV, HCV), or severe uncontrolled chronic disease.

LJI participants received an intramuscular late-season priming dose of standard-dose quadrivalent inactivated influenza vaccine (Fluzone 2023–2024 formulation, Sanofi Pasteur) in May–June 2024. Four months later (September-November 2024), 14 participants returned to receive a booster dose of standard-dose trivalent inactivated influenza vaccine (Fluzone 2024–2025, Sanofi Pasteur). Serum was collected at day 0 (pre-prime, May/June 2024), day 28 (July 2024), day 90 (September 2024), day 120 (pre-boost, September-November 2024), day 150 (28 days post-boost, December 2024), and day 210 (90 days post-boost, February 2025).

To benchmark late-season prime-boost dynamics against standard annual regimens, we evaluated an independent cohort from the University of Georgia (UGA cohort). Forty-one healthy participants aged 18–89 received standard fall influenza vaccinations in both 2023–2024 and 2024–2025. UGA participants under 65 years old received standard-dose Fluzone Quadrivalent, while participants aged 65 or older received Fluzone High-Dose (Sanofi Pasteur). Sera were collected at days 0, 30, and 90 post-2023 vaccination as well as days 365 and 395 post-prime the following year (days 0 and 30 post-2024 vaccination).

Current influenza vaccination guidelines allow one influenza vaccine to be received each season, with the season ending each June 30. Thus, the 2023–2024 vaccine can be given from July 1, 2023 until June 30, 2024 while the 2024–2025 vaccine can be given from July 1, 2024 until June 30, 2025. Thus, to assess prime-boost influenza vaccinations, we administered the 2023 vaccine in May 2024 (at the end of the 2023 season) to individuals that had not been vaccinated the prior fall, with the booster vaccine administered the following fall to match standard vaccine timing.

### Influenza viruses

Serum antibody titers were quantified using HAI assays against a panel of 17 influenza variants across four subtypes/lineages: H1N1, H3N2, B/Victoria, and B/Yamagata (**Table 2**). All viral strains were propagated in embryonated chicken eggs, with the exception of H3N2 A/Massachusetts/18/2022, which was cell-grown in MDCK cells.

For the LJI cohort, HAI titers were evaluated across all 17 strains at every sampling time point. For the UGA cohort, 14 of the 17 strains were measured at all time points. Specific exclusions in the UGA dataset included H3N2 A/Hong Kong/4801/2014 and A/Massachusetts/18/2022 (not evaluated at days 0, 30, and 90 post-2023 vaccination) and H1N1 A/Victoria/4897/2022 (not evaluated at day 90 post-2023 vaccination).

### Hemagglutination inhibition (HAI) protocol

For the UGA cohort, sera were treated with receptor-destroying enzyme (RDE, Denka) to remove non-specific inhibitors. Briefly, three volumes of RDE were added to one volume of serum and incubated overnight at 37°C, followed by heat inactivation at 56°C for 30–60 minutes and dilution with 6 volumes of 1x phosphate-buffered saline (PBS) to achieve a starting dilution of 1:10. For the LJI cohort, 20 µL of serum was treated with 10 µL of 8 mg/mL TPCK-trypsin (Sigma-Aldrich) at 56°C for 30 minutes, followed by the addition of 60 µL of 11 mM potassium periodate (Sigma-Aldrich) for 15 minutes at room temperature, neutralization with 60 µL of 1% glycerol-PBS for 15 minutes, and dilution with 50 µL of 0.85% PBS.

Sera were serially diluted 2-fold in V-bottom 96-well plates. An equal volume of influenza virus standardized to 8 haemagglutination units (HAU)/50 µL was added to each well. Plates were incubated at room temperature for 20 minutes (UGA) or 30 minutes (LJI). Subsequently, 50 µL of 0.8% turkey red blood cells (Lampire Biologicals, Pipersville, PA, USA) was added to all wells, except for H3N2 A/Darwin/9/2021 in the LJI cohort, which utilized 0.75% guinea pig red blood cells due to reduced viral agglutination with turkey RBCs. Plates were incubated for 30 minutes at room temperature (turkey RBCs) or 45 minutes at 4°C (guinea pig RBCs) prior to reading.

The HAI titer was defined as the reciprocal of the highest serum dilution producing complete inhibition of haemagglutination. Positive control sera and negative control wells were included on every plate to confirm assay consistency.

Throughout this work, non-responders to a vaccine strain were defined as subjects with HAI≤10 at all time points against that strain.

### Statistical analysis

Statistical significance was defined as p≤0.05 in all analyses and denoted by an asterisk in figures. Exact *p*-values are reported for all statistical testing, with values less than 0.001 reported as p<0.001 for simplicity. Differences in HAI titers between prespecified time points or vaccine strains were assessed using Welch’s one-sided t-tests on log_2_-transformed HAI titers.To account for multiple testing, raw *p*-values were adjusted using the Holm method for each influenza vaccine strain to control the family-wise error rate.

The antibody ceiling effect was computed using the geometric mean titers across all variants within a subtype or lineage. Taking H1N1 as an example, the geometric mean titer across all variants at day 0 (GMT_0_) and at day 30 (GMT_30_) were computed for each subject, and the resulting points (GMT_0_, GMT_30_/GMT_0_) were fit using linear regression to infer the slope that defines the ceiling effect.

To quantify study-study similarity, RMSE was computed between the log_2_(HAI/5) of two individuals across all measured influenza variants for H1N1, H3N2, or B/Victoria. The results were then exponentiated by 2 to convert into un-logged units. The number of pairs of subjects with (pre- and post-vaccination RMSE≤4x) divided by the number of pairs with (pre-vac RMSE≤4x and post-vaccination RMSE>4x) quantified study similarity. For cross-study similarity, one subject from each pair must have come from each study. For within-study similarity, both pairs of subjects came from the same study.

## Data Availability

Upon publication, all serological data and metadata will be made available with the manuscript.

## Acknowledgements

We are deeply grateful to the La Jolla Institute for Immunology (LJI) Clinical Core and the University of Georgia (UGA) FRIC influenza vaccine teams for donor enrollment, sample collection, blood processing, and technical assistance, and we acknowledge Julia Aguirre, Brittany Baker, Brianna Gamez, Hannah B. Hanley, Erin Jarrett, Hana Ji, Maya McCoy, Zachary McGuire, Frankie Stewart, Naoko Uno, and Emma Whitesell. In addition, we would like to thank all the participants for their donations and time in contributing samples to this study. Certain influenza A and B viruses were obtained through the Prevention and Control of Influenza, Centers for Disease Control and Prevention, Atlanta, GA, USA and the International Reagent Resource.

## Supplementary Figures and Tables

**Figure S1.**
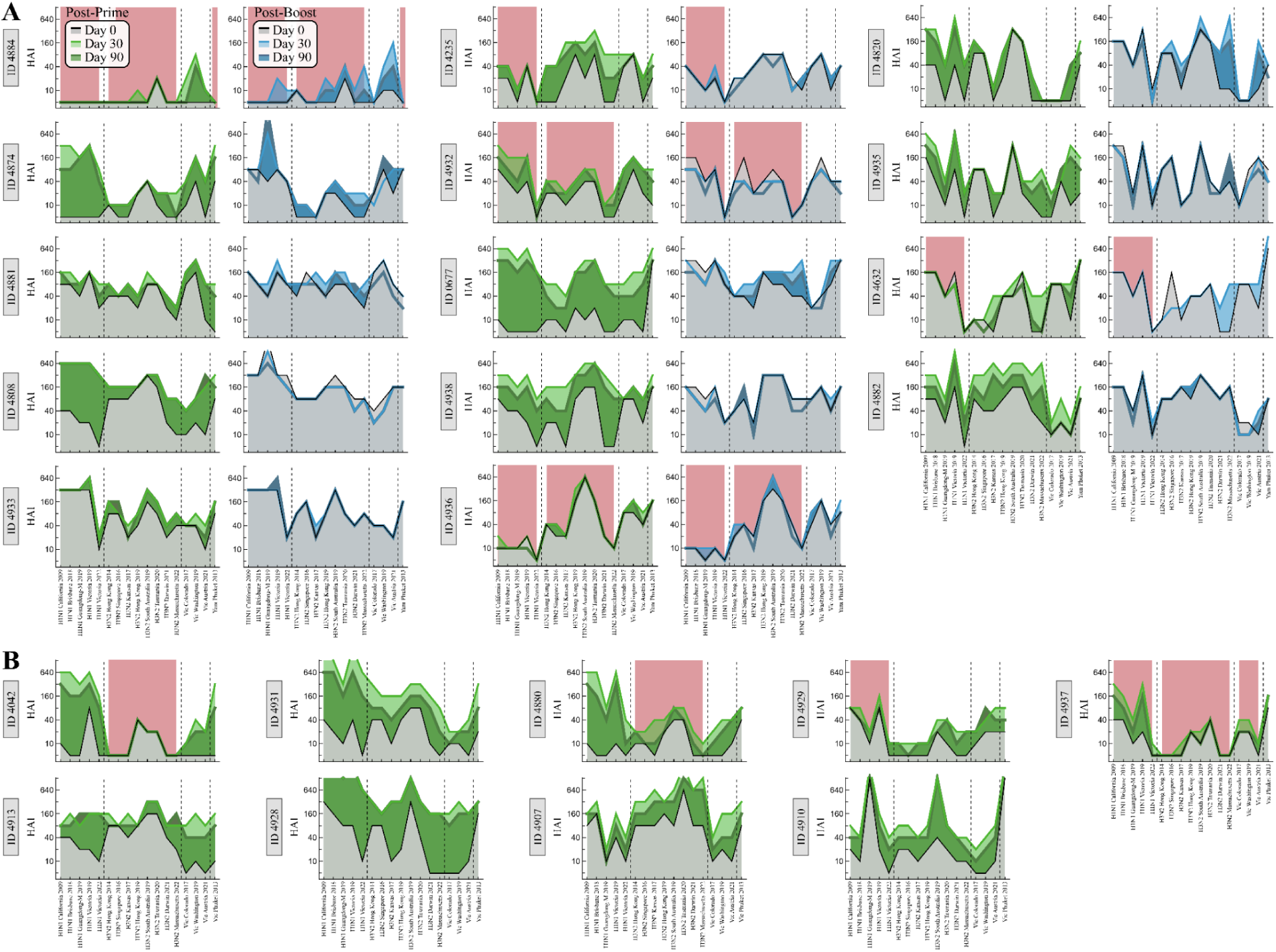
Individual antibody landscapes for the LJI cohort. Titers at all time points from subjects that (A) received the prime and booster vaccines or (B) only received the priming vaccine. In each panel, subjects are sorted from least-to-greatest day 0 GMT across all variants. H1N1, H3N2, B/Victoria, or B/Yamagata strains have a red highlight for subjects with HAI titers≤10 for all time points for that vaccine strain (with either H3N2 vaccine strain counting).

**Figure S2.**
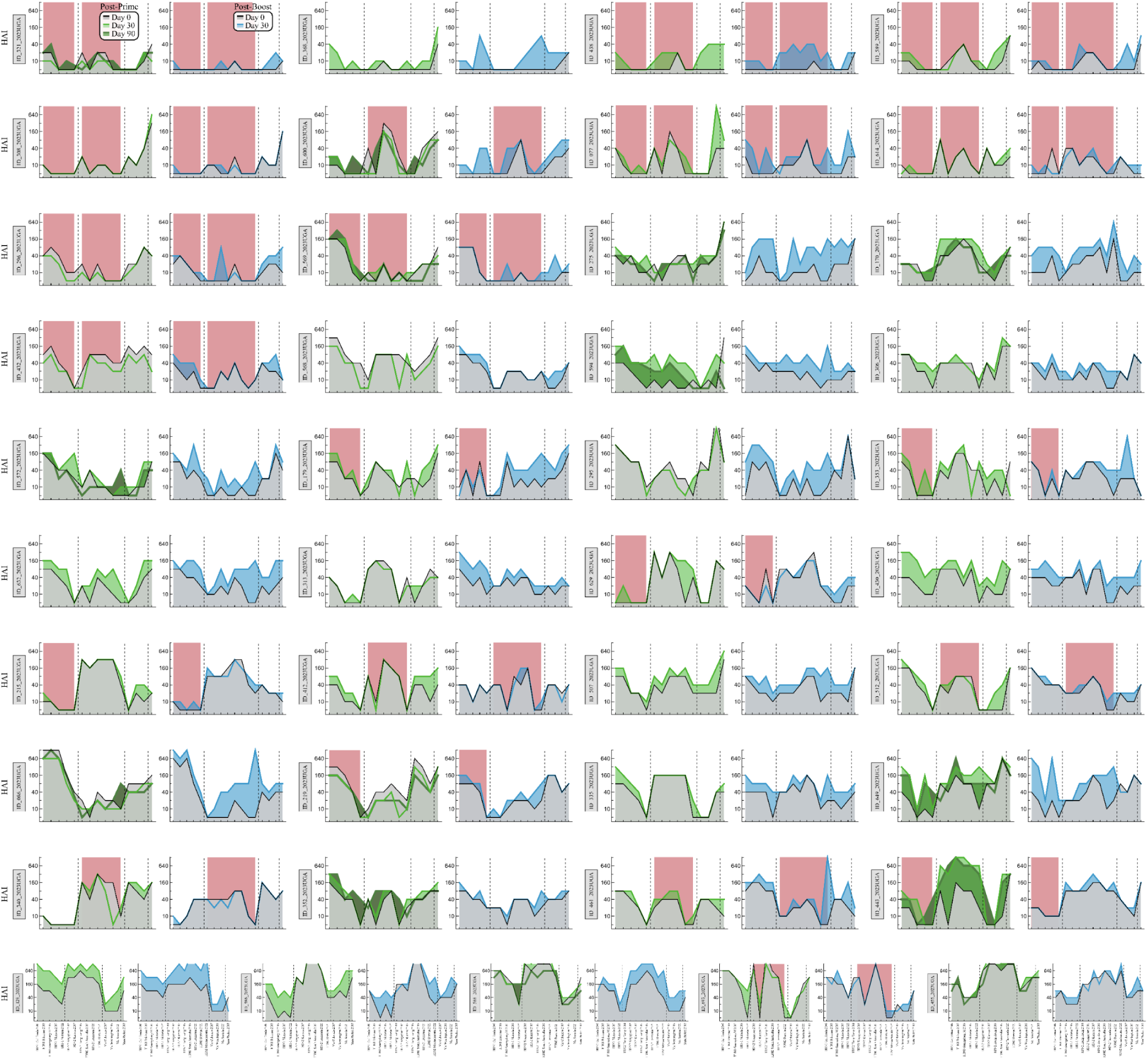
Individual antibody landscapes for the UGA cohort. Titers at all time points from UGA subjects receiving a vaccine in the fall of 2023 and the fall of 2024. Subjects are sorted from least-to-greatest day 0 GMT across all variants. The 2023 virus panel did not include H3N2 Hong Kong 2014 or Massachusetts 2022, and H1N1 Victoria 2022 was not measured at day 90. H1N1, H3N2, B/Victoria, or B/Yamagata strains have a red highlight for subjects with HAI titers≤10 for all time points for that vaccine strain (with either H3N2 vaccine strain counting).

**Figure S3.**
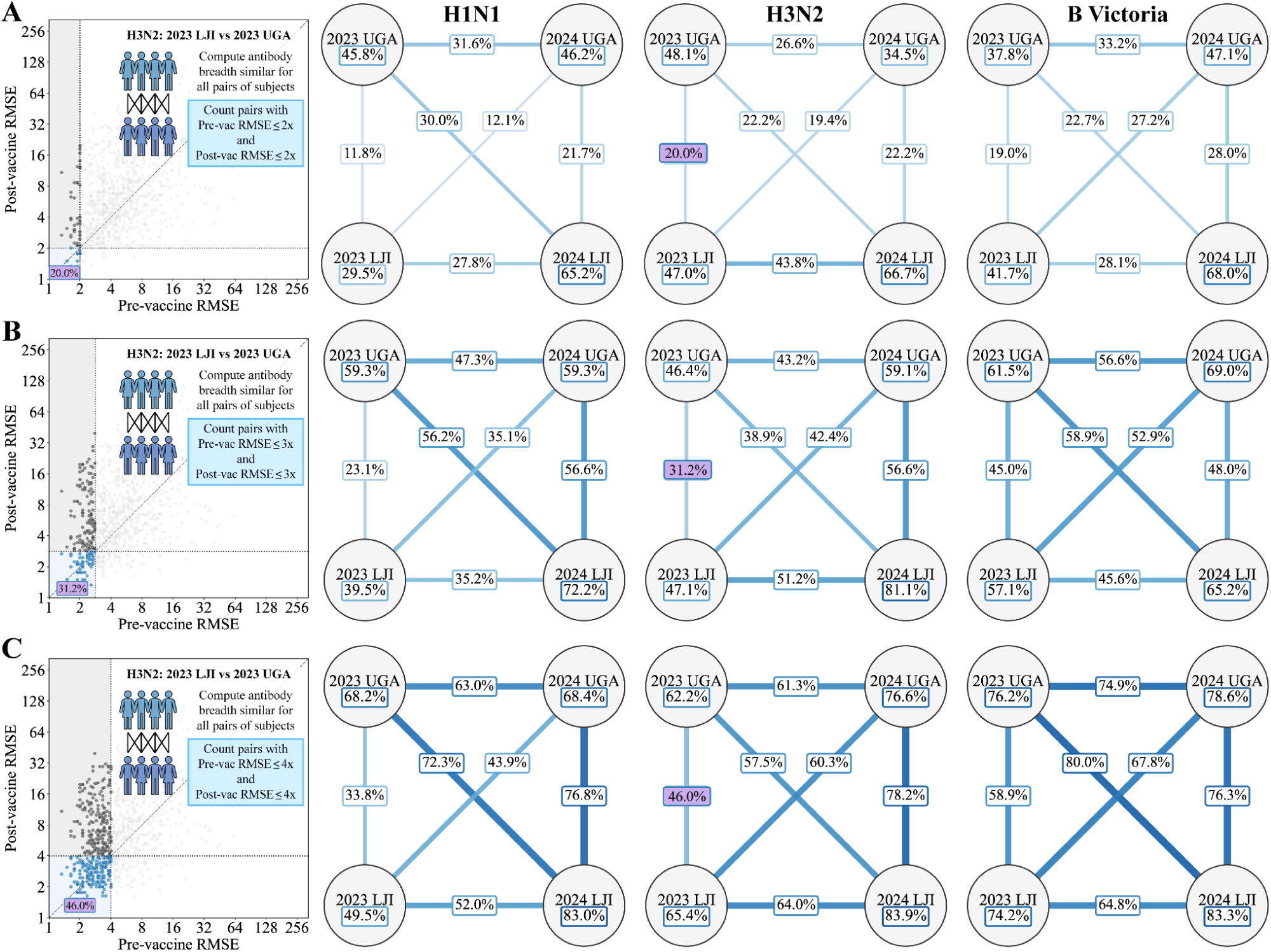
RMSE-based matching of antibody landscapes between LJI and UGA study cohorts. Pre- and post-vaccination RMSE distribution for all 2023 UGA and 2023 LJI subject pairs against H3N2 variants, with the purple region indicating pairs meeting the similarity threshold of (A) RMSE≤2x, (B) ≤3x, and (C) ≤4x. Each threshold’s corresponding cohort similarity networks for H1N1, H3N2, and B/Victoria are shown to the left. Connection labels indicate the fraction of subject pairs exhibiting pre- and post-vaccine RMSE less than or equal to each threshold across internal cohort comparisons and cross-cohort alignments. B/Yamagata was excluded because only a single variant was measured from that lineage.

**Table S1.**
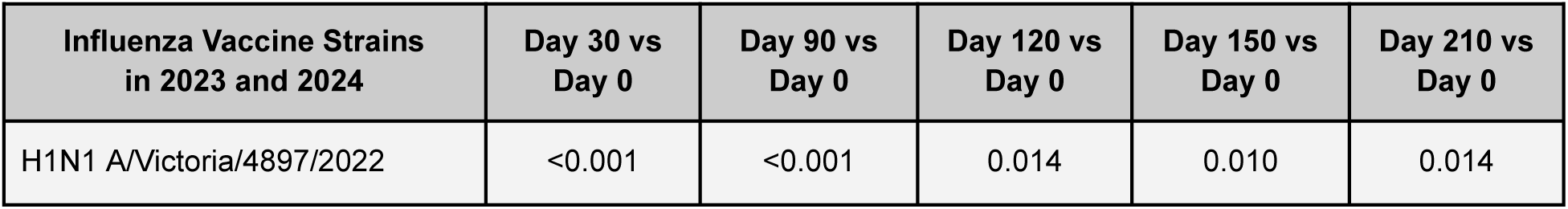
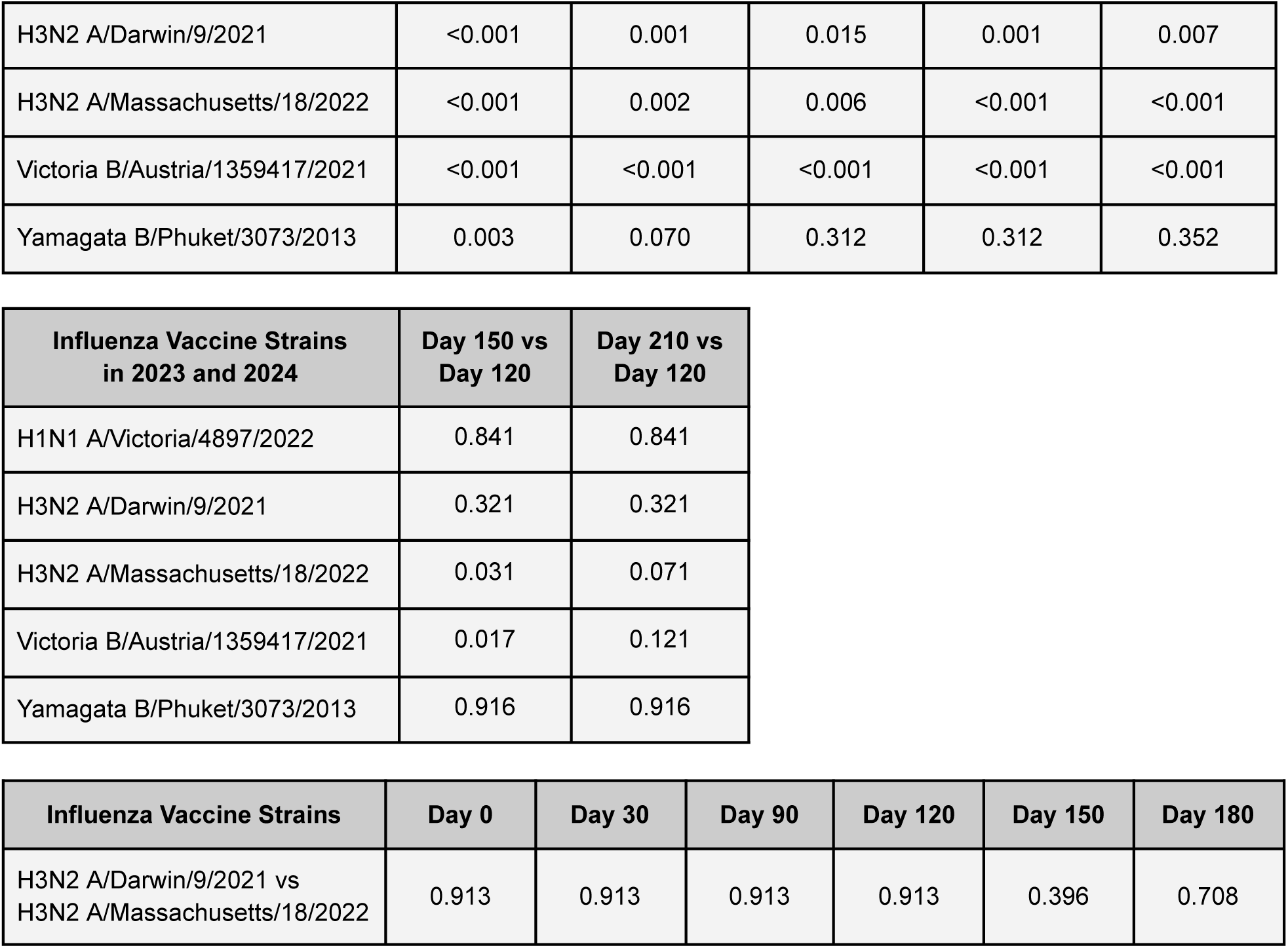
Statistical significance of HAI titer increases at each time point compared to day 0, day 120, or between the 2023-2024 H3N2 vaccine strains in the LJI cohort. *p*-values were based on one-sided t-test with Holm correction applied for each influenza vaccine strain. When comparing the 2023-2024 H3N2 vaccine strains, the strain in the most recently administered vaccine was expected to have the higher titer (*i.e.*, H3N2 A/Darwin/9/2021 at days 0, 30, and 90; H3N2 A/Massachusetts/18/2022 at days 120, 150, and 210).

| Influenza Vaccine Strains in 2023 and 2024 | Day 30 vs Day 0 | Day 90 vs Day 0 | Day 120 vs Day 0 | Day 150 vs Day 0 | Day 210 vs Day 0 |
| --- | --- | --- | --- | --- | --- |
| H1N1 A/Victoria/4897/2022 | <0.001 | <0.001 | 0.014 | 0.010 | 0.014 |
| H3N2 A/Darwin/9/2021 | <0.001 | 0.001 | 0.015 | 0.001 | 0.007 |
| H3N2 A/Massachusetts/18/2022 | <0.001 | 0.002 | 0.006 | <0.001 | <0.001 |
| Victoria B/Austria/1359417/2021 | <0.001 | <0.001 | <0.001 | <0.001 | <0.001 |
| Yamagata B/Phuket/3073/2013 | 0.003 | 0.070 | 0.312 | 0.312 | 0.352 |

| Influenza Vaccine Strains in 2023 and 2024 | Day 150 vs Day 120 | Day 210 vs Day 120 |
| --- | --- | --- |
| H1N1 A/Victoria/4897/2022 | 0.841 | 0.841 |
| H3N2 A/Darwin/9/2021 | 0.321 | 0.321 |
| H3N2 A/Massachusetts/18/2022 | 0.031 | 0.071 |
| Victoria B/Austria/1359417/2021 | 0.017 | 0.121 |
| Yamagata B/Phuket/3073/2013 | 0.916 | 0.916 |

| Influenza Vaccine Strains | Day 0 | Day 30 | Day 90 | Day 120 | Day 150 | Day 180 |
| --- | --- | --- | --- | --- | --- | --- |
| H3N2 A/Darwin/9/2021 vs H3N2 A/Massachusetts/18/2022 | 0.913 | 0.913 | 0.913 | 0.913 | 0.396 | 0.708 |

**Table S2.** Statistical significance of HAI titer increases at each time point compared to day 0, day 365, or between the 2023-2024 H3N2 vaccine strains in the UGA cohort. *p*-values based on one sided t-test, with Holm correction applied for each influenza vaccine strain. Unlike in the LJI cohort, day 90 titers were not measured for H1N1, and since the H3N2 A/Massachusetts/18/2022 was not measured at day 0, 30, or 90. When comparing the 2023-2024 H3N2 vaccine strains at days 365 and 395, H3N2 A/Massachusetts/18/2022 was expected to have the higher titers.

| Influenza Vaccine Strains in 2023 and 2024 | Day 30 vs Day 0 | Day 90 vs Day 0 | Day 365 vs Day 0 | Day 395 vs Day 0 |
| --- | --- | --- | --- | --- |
| H1N1 A/Victoria/4897/2022 | 0.040 | N/A | 0.019 | <0.001 |
| H3N2 A/Darwin/9/2021 | 0.071 | 0.191 | 0.898 | 0.071 |
| Victoria B/Austria/1359417/2021 | 0.006 | 0.266 | 0.964 | 0.266 |
| Yamagata B/Phuket/3073/2013 | 1 | 1 | 1 | 1 |

| Influenza Vaccine Strains in 2023 and 2024 | Day 395 vs Day 365 |
| --- | --- |
| H1N1 A/Victoria/4897/2022 | <0.001 |
| H3N2 A/Darwin/9/2021 | <0.001 |
| H3N2 A/Massachusetts/18/2022 | <0.001 |
| Victoria B/Austria/1359417/2021 | <0.001 |
| Yamagata B/Phuket/3073/2013 | 0.074 |

| Influenza Vaccine Strains | Day 365 | Day 395 |
| --- | --- | --- |
| H3N2 A/Darwin/9/2021 vs H3N2 A/Massachusetts/18/2022 | 0.283 | 0.011 |

